## Supplementary Figures and Tables for "PD-L1 mediated T cell inhibition by regulatory plasma cells induced after sepsis and COVID-19"

### Supplementary Material

**Fig S1. Innate immune cells in spleen of Sham and CLP mice.**

(A) Proportions among splenocytes and absolute counts of dendritic cells in 7 Sham and 12 CLP mice. (B) Proportions among splenocytes and absolute counts of NK cells in 13 Sham and 13 CLP mice. (C) Proportions among splenocytes and absolute counts of CD11b+ myeloid cells in 10 Sham and 13 CLP mice. (D) Proportions among CD11b+ myeloid cells and absolute counts of monocytes/macrophages in 10 Sham and 13 CLP mice. (E) Proportions among CD11b+ myeloid cells and absolute counts of neutrophils in 10 Sham and 13 CLP mice.

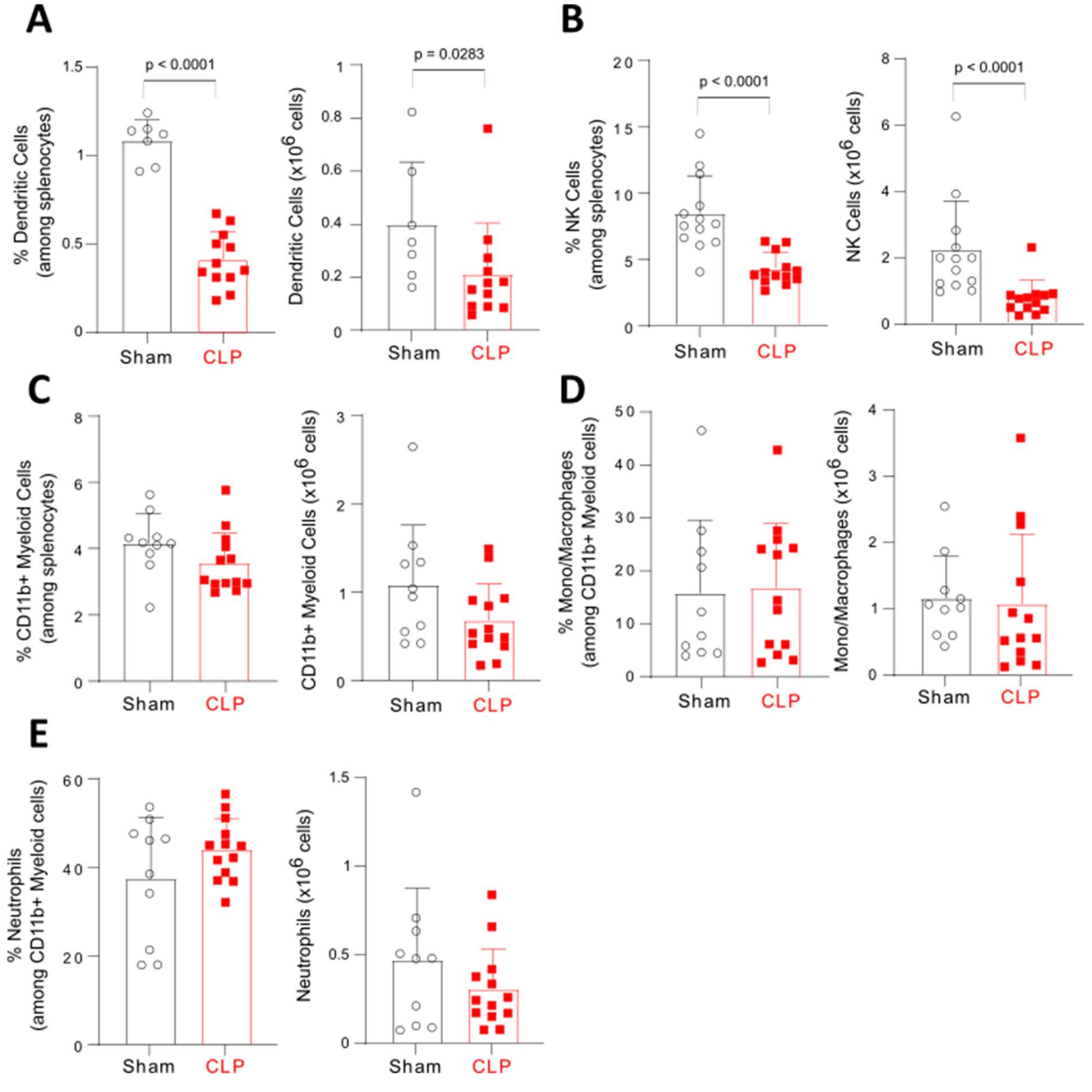

**Fig S2. T lymphocyte subpopulations in spleen of Sham and CLP mice.**

(A) Proportions of CD4+ and CD8+ T cells among splenocytes in 23 Sham and 25 CLP. (B-D) Proportions among CD4+ T cells and absolute counts of naïve, central memory and effector CD4+ T lymphocytes in 11 Sham and 13 CLP mice. (E-G) Proportions among CD8+ T cells and absolute counts of naïve, central memory and effector CD8+ T lymphocytes in 11 Sham and 13 CLP mice.

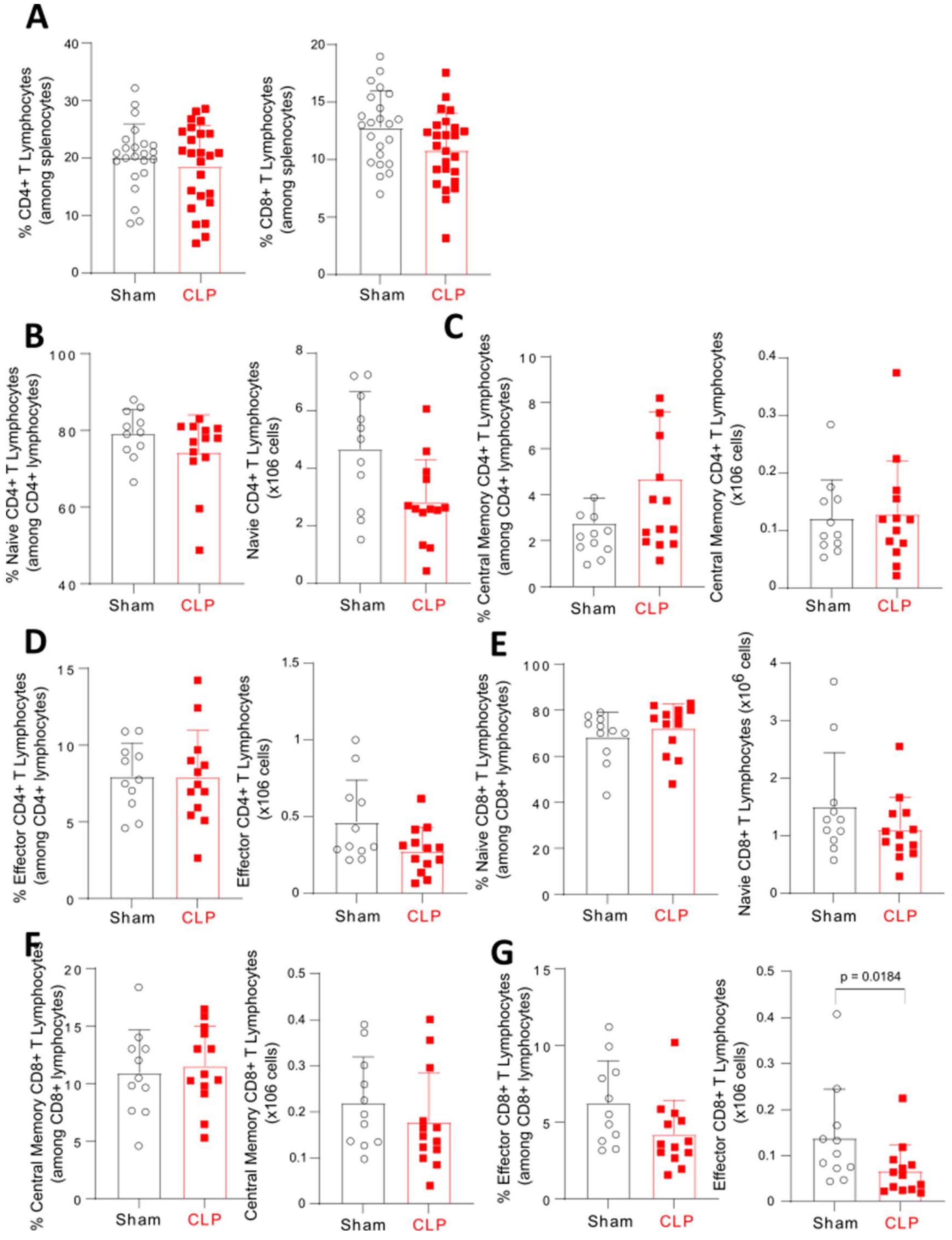

**Fig S3. Proliferation capacity of splenic T cells from Sham and CLP mice.**

(A-B) Splenocytes from 5 Sham and 5 CLP mice were stimulated *ex vivo* by anti-CD3/CD28 Abs-coated beads. Cell proliferation was measured among viable cells by flow cytometry. Percentages of proliferating CD4+ (A) and CD8+ (B) T cells were evaluated. (C) Splenocytes from 5 Sham and 5 CLP mice were stimulated *ex vivo* by phytohemagglutinin. Cell proliferation was measured among viable cells by flow cytometry. Total percentages of proliferating CD3+ T cell and percentages of proliferating CD3+ T cell undergoing more than 3 or 4 division cycles were evaluated. Red squares represent results in CLP mice and white circles in Sham animals. Geometric means with standard deviations (SD) and individual values are shown. Mann-Whitney tests were performed and only significant p values < 0.05 are shown.

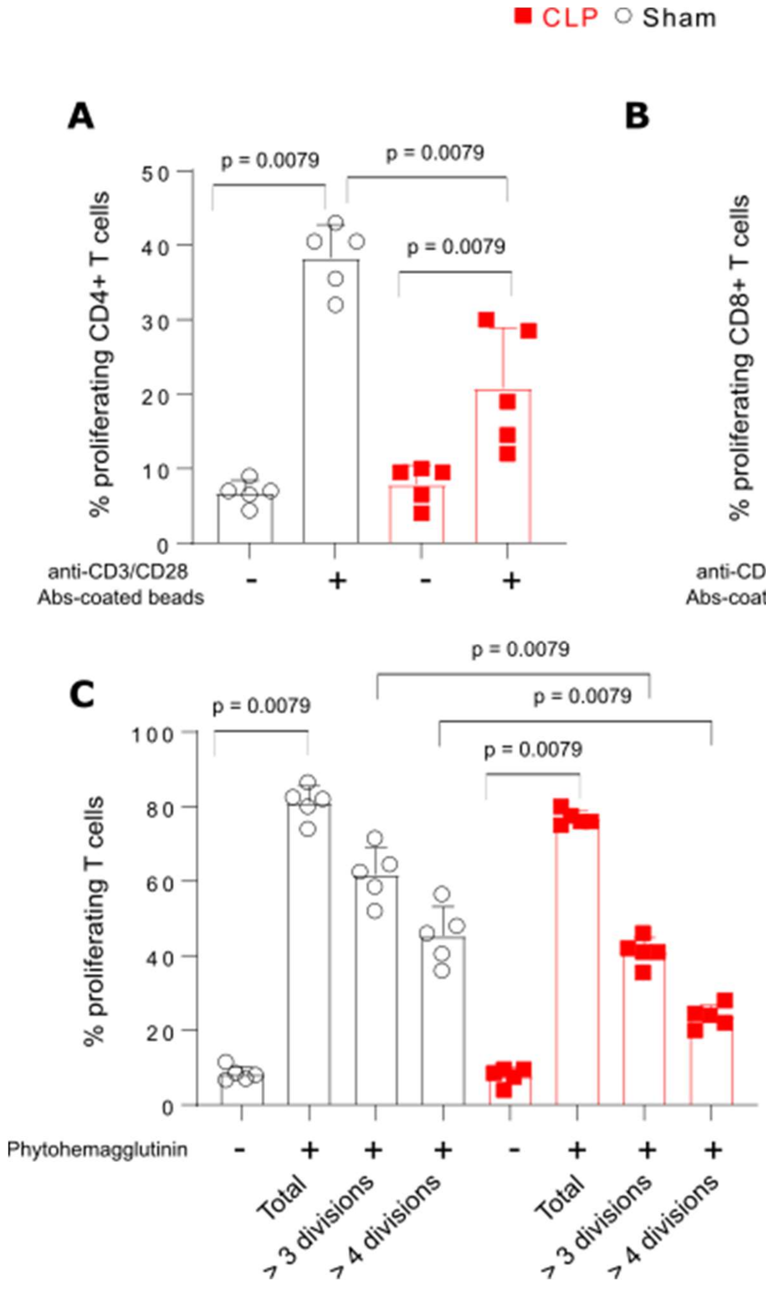

**Fig S4. Impact of the addition of B cells and plasma cells from septic patients in T cell co-culture assay.**

Purified T cells from blood of healthy donors (n = 8) were cultured *ex vivo* in the absence or in the presence of stimulating anti-CD2/CD3/CD28 antibodies-coated beads, either alone or with B cells (n = 6) or plasma cells (n = 8) from septic patients at 1:1 ratio in the stimulated condition. Proportions of proliferating CD4+ (A) and CD8+ (B) T cells were measured by flow cytometry among viable CD4+ and CD8+ T cells respectively. Box-plots and individual values are shown. Non-parametric Wilcoxon paired tests were performed to compare results from different co-culture conditions. Only significant p values < 0.05 are shown.

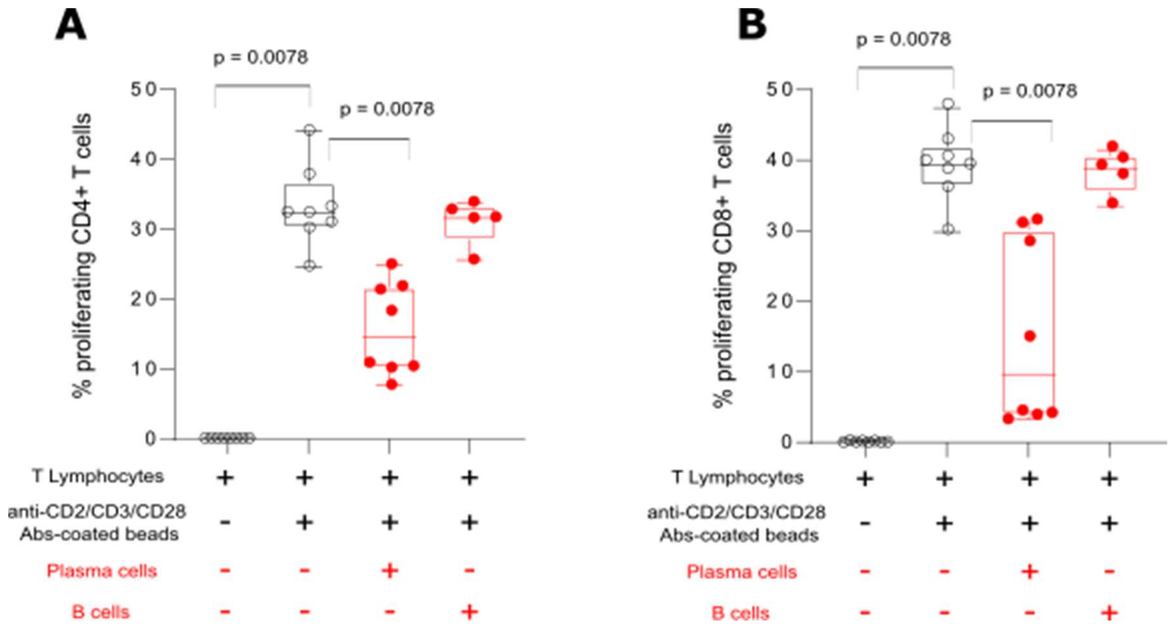

**Fig S5. Flow cytometry gating strategy and representative examples in mice for cell surface stainings on B cells and plasma cells.**

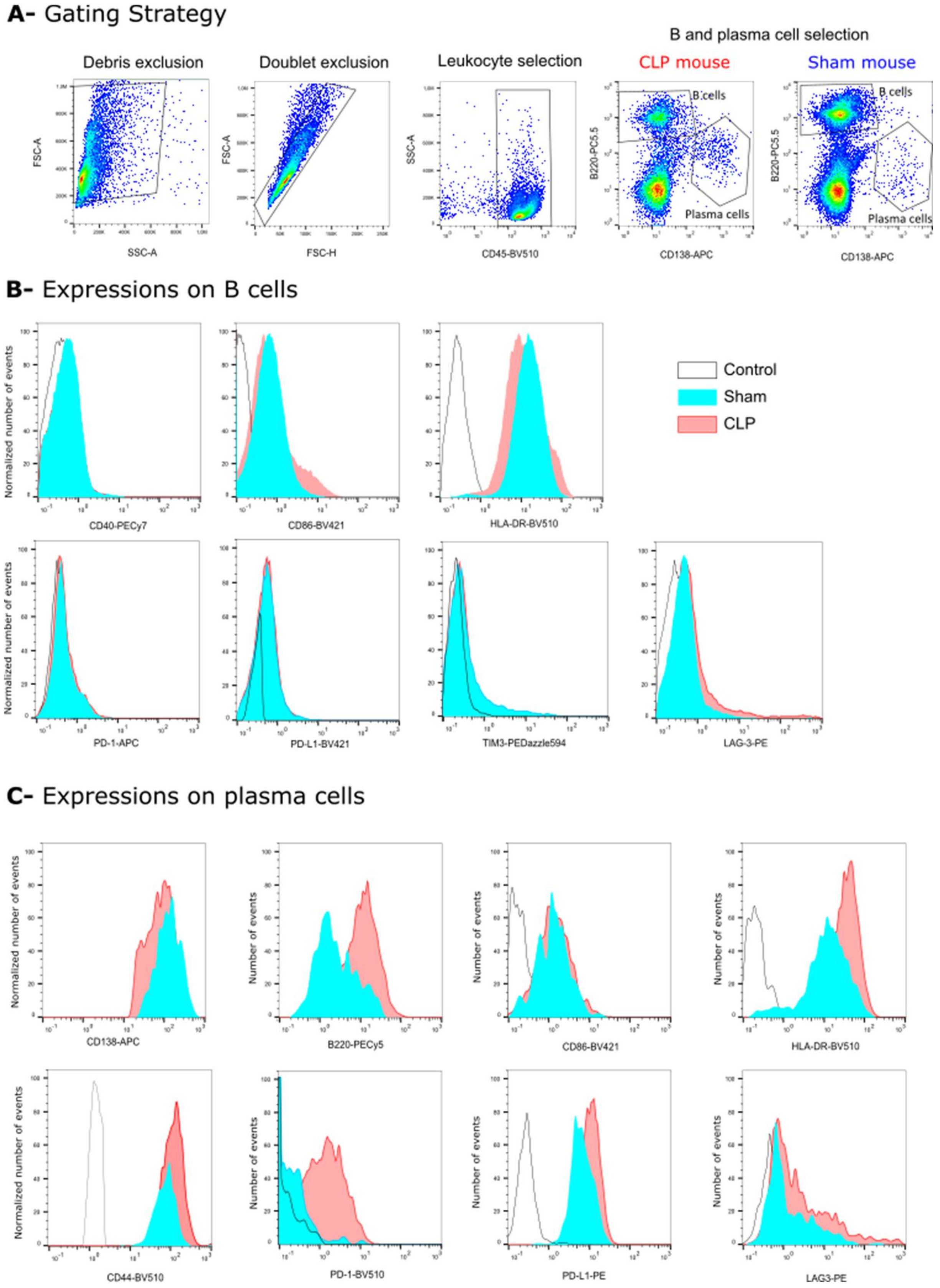

**Fig S6. Flow cytometry gating strategy and representative example of BLIMP-1 intracellular stainings in B cells and plasma cells from one CLP mouse.**

**A-** Gating Strategy

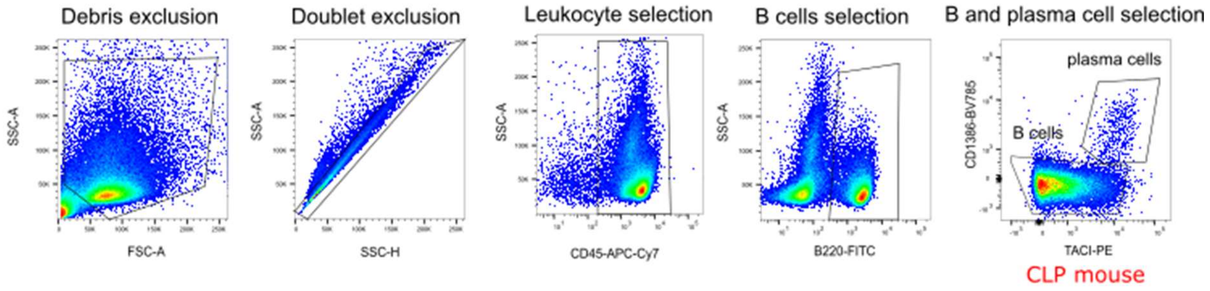

**B-** Expressions on B cells and plasma cells in CLP mice

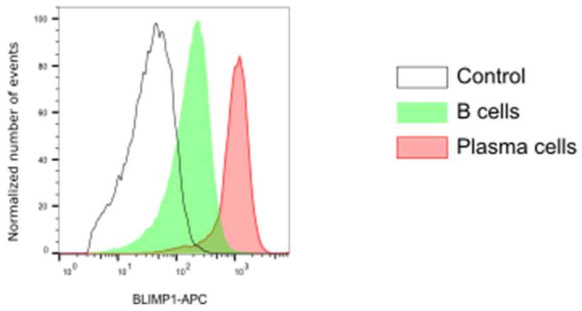

**Fig S7. Flow cytometry gating strategy and representative examples in healthy donors and patients for cell surface and intracellular stainings.**

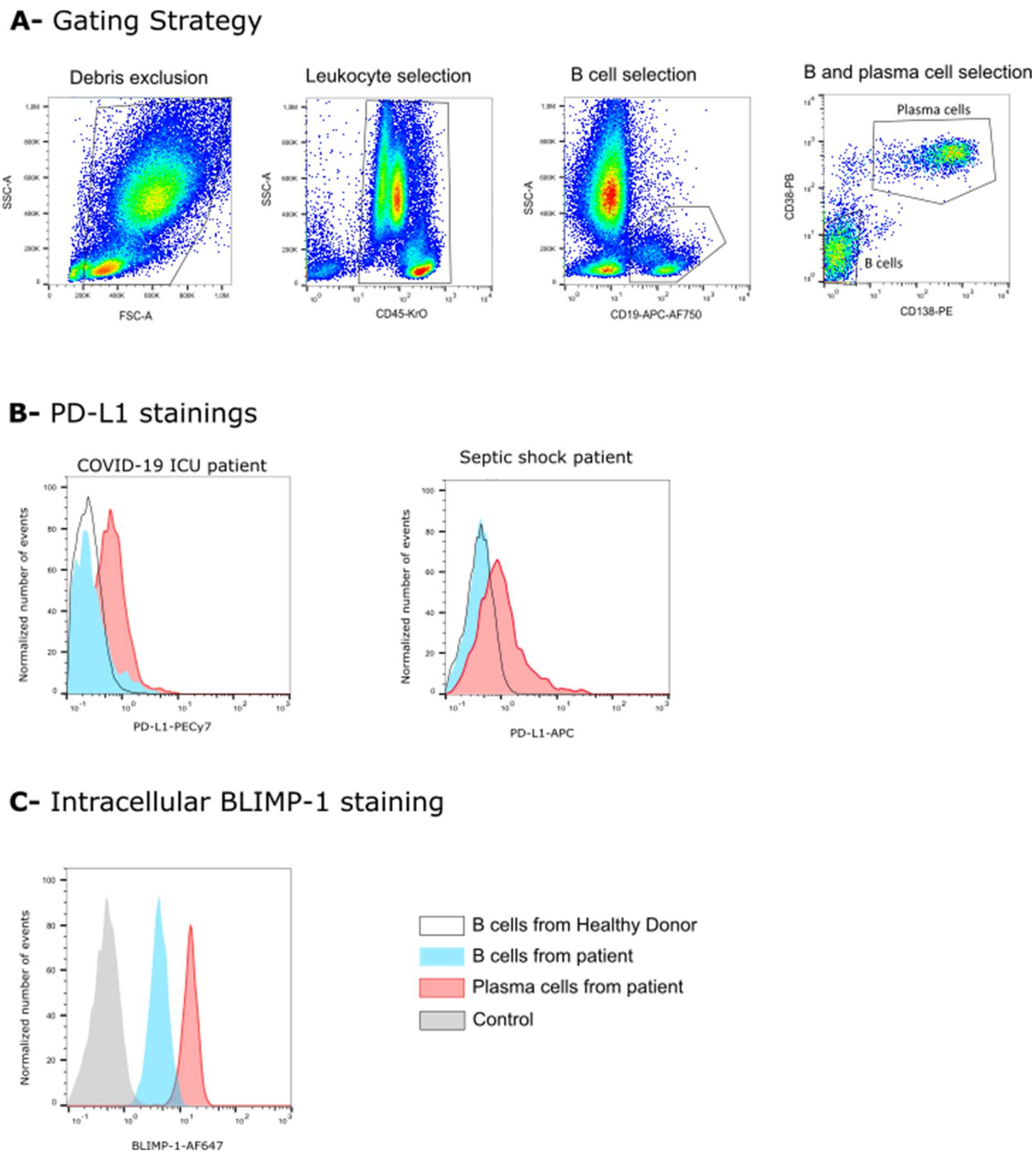

**Table S1. Demographic and clinical data for critically ill patients with bacterial sepsis and COVID-19.** Values are presented as numbers and percentages for categorical variables and medians and [Q1-Q3] interquartiles ranges for continuous variables. SOFA: Sepsis-related organ failure assessment score. SAPS II: simplified acute physiology score. BGP: bacilli Gram negative. CGP: cocci Gram positive. mHLA-DR: MHC class II expression on monocytes. AB/C: numbers of antibodies bound per cells. Normal values = 13 500 – 45 000 AB/C. Age-matched normal values of CD4$^{+}$ T cell count = 336 – 1126 cell/µL.

| | REALISM<br>n= 107 | IMMUNOSEPSIS<br>n = 67 | RICO<br>n = 33 |
|---|---|---|---|
| Type of study | Retrospective | Prospective | Prospective |
| Age | 68 [59 – 77] | 72 [61 – 78] | 65 [52 – 70] |
| Gender (Male) | 69 (64.5) | 42 (63) | 25 (76) |
| MacCabe score<br>0<br>1<br>2 | <br>66 (62)<br>34 (32)<br>7 (6) | <br>38 (57)<br>27 (40)<br>2 (3) | <br>29 (88)<br>3 (9)<br>1 (3) |
| Charlson score<br>0<br>≥1 | <br>26 (24)<br>81 (76) | <br>12 (18)<br>55 (82) | <br>16 (49)<br>17 (52) |
| SOFA score | 8.5 [7 – 11]<br>** 15 missing values* | 9 [7 – 11] | 3 [2 – 6] |
| SAPS II score | 47 [37 – 55] | 54 [42 – 69] | 32 [26 – 41] |
| Infection type | bacterial sepsis | bacterial sepsis | viral sepsis |
| Infection diagnosis<br>Microbiology<br>Radiology<br>Surgery<br>Suspected | <br>79 (74)<br>11 (10)<br>8 (8)<br>9 (8) | *1 missing value*<br>45 (68)<br>10 (15)<br>5 (8)<br>6 (9) | <br>33 (100)<br>0<br>0<br>0 |
| Site of infection<br>Pulmonary<br>Abdominal<br>Other | <br>29 (28)<br>43 (41)<br>31 (30) | *3 missing values*<br>12 (19)<br>27 (42)<br>25 (39) | <br>33 (100)<br>0<br>0 |
| Pathogen type (% among microbiologically proven infections)<br>BGN<br>CGP<br>Fungi<br>Virus<br>Other<br>Unidentified | <br>31 (39)<br>32 (41)<br>10 (13)<br>2 (3)<br>5 (6)<br>16 (20) | <br>30 (67)<br>18 (40)<br>3 (7)<br>0<br>2 (4)<br>0 | <br>0<br>0<br>0<br>33 (100)<br>0<br>0 |
| Mortality at D14 | 14 (13) | 9 (13) | 2 (6) |
| ICU-acquired infections | 20 (19) | 10 (15) | 15 (46) |
| Immune status at D3<br>mHLA-DR (AB/C)<br><br>CD4+ T cells (cells/µL) | <br>5 565 [3 178 – 9 546]<br>** 19 missing values*<br>397 [280 – 651]<br>** 19 missing values* | <br>5 012 [3 308 – 8 588]<br><br>350 [197 – 557] | <br>8 288 [5 868 – 10 808]<br>** 9 missing values*<br>348 [223 – 412]<br>** 9 missing values* |

**Table S2. Severity score criteria for CLP model evaluation**

| | | | | |
|---|---|---|---|---|
| **Fur aspect** | Active grooming | Succinct grooming | Dull coat or bristly hair | Piloerection |
| **Motor activity** | Normal | Decreased but reactive if stimulated | Decreased even if stimulated | None |
| **Posture** | Normal | Arched back | Prostrate but free movements | Prostrate motionless, twisted body |
| **Breathing** | Normal | Normal | Moderate dyspnea | Severe dyspnea |
| **Weight loss** | 0 - 5 % | 5 – 10 % | 10 – 15 % | > 15 % |
| **Score** | 0 | 1 | 2 | 3 |

### Table S3. Key Resources Table

| REAGENT or RESOURCE | SOURCE | IDENTIFIER | LOT NUMBER |
|---|---|---|---|
| Antibodies | | | |
| Rat anti-mouse Blimp-1 (APC) (clone 5E7) | BioLegend | Cat#150008, RRID:AB_2728187 | B321722 |
| Rat anti-mouse CD1d (AF647) (clone 1B1) | BioLegend | Cat#123512, RRID:AB_1236532 | B194780 |
| Armenian Hamster anti-mouse CD3ε (PE) (clone 145-2C11) | BioLegend | Cat#100308, RRID:AB_312673 | B376518 |
| Rat anti-mouse CD3 (BV785) (clone 17A2) | BioLegend | Cat#100231, RRID:AB_11218805 | B365876 |
| Rat anti-mouse CD4 (BV421) (clone GK1.5) | BioLegend | Cat#100443, RRID:AB_2562557 | B213276; B275080 |
| Rat anti-mouse CD4 (APC) (clone GK1.5) | BioLegend | Cat#100412, RRID:AB_312696 | B372226 |
| Rat anti-mouse CD5 (BV421) (clone 53-7.3) | BioLegend | Cat#100617, RRID:AB_2562173 | B208111; B213474 |
| Rat anti-mouse CD8a (PE-Cy7) (clone 53-6.7) | BioLegend | Cat#100722, RRID:AB_312761 | B222017 |
| Rat anti-mouse/human CD11b (BV421) (clone M1/70) | BioLegend | Cat#101251, RRID: AB_11203704 | B212710 |
| Rat anti-mouse/human CD11b (FITC) (clone M1/70) | BioLegend | Cat#101206, RRID: AB_312789 | B349919 |
| Armenian Hamster anti-mouse CD11c (APC) (clone N418) | BioLegend | Cat#117310, RRID: AB_313779 | B206713 |
| Rat anti-mouse CD25 (APC) (clone 3C7) | BioLegend | Cat#101910, RRID:AB_2280288 | B218018 |
| Rat anti-mouse CD40 (PE-Cy7) (clone 3/23) | BioLegend | Cat#124622, RRID:AB_10897812 | B226722 |
| Rat anti-mouse CD44 (BV421) (clone IM7) | BioLegend | Cat#103040, RRID:AB_2616903 | B228179 |
| Rat anti-mouse/human CD44 (BV711) (clone IM7) | BioLegend | Cat#103057, RRID:AB_2564214 | B384036 |
| Rat anti-mouse CD45 (APC-Fire 750) (clone 30-F11) | BioLegend | Cat#103154, RRID:AB_2572116 | B226658; B250950; B260280 |

| | | | |
|---|---|---|---|
| Rat anti-mouse CD45 (BV510) (clone 30-F11) | BioLegend | Cat#103138, RRID:AB_2563061 | B220123; B220124; B232692; B235434; B240739; B251566; B265567; B280070; B280079; B386738 |
| Rat anti-mouse/human CD45R/B220 (FITC) (clone RA3-6B2) | BioLegend | Cat#103206, RRID:AB_312991 | B230445; B247731 |
| Rat anti-mouse/human CD45R/B220 (PE-Cy7) (clone RA3-6B2) | BioLegend | Cat#103222, RRID:AB_313005 | B246946; B256217 |
| Rat anti-mouse CD49B (APC) (clone DX5) | BioLegend | Cat#108910, RRID: AB_313417 | B375231 |
| Rat anti-mouse CD62L (BV421) (clone MEL-14) | BioLegend | Cat#104435, RRID:AB_2562560 | B372292 |
| Armenian Hamster anti-mouse CD80 (APC) (clone 16-10A1) | BioLegend | Cat#104714, RRID:AB_313135 | B227918 |
| Rat anti-mouse CD86 (BV421) (clone GL-1) | BioLegend | Cat#105031, RRID:AB_10898329 | B223811 |
| Rat anti-mouse CD115 (PE) (clone AFS98) | BioLegend | Cat#135506, RRID:AB_1937253 | B190335; B190336 |
| Rat anti-mouse CD127 (PE) (clone A7R34) | BioLegend | Cat#135010, RRID:AB_1937251 | B185924; B185925 |
| Rat anti-mouse CD138 (APC) (clone 281-2) | BioLegend | Cat#142506, RRID:AB_10962911 | B197972; B237677; B239451; B270362; |
| Rat anti-mouse CD138 (BV785) (clone 281-2) | BioLegend | Cat#142534, RRID:AB_2814047 | B366379 |
| Rat anti-mouse CD223 (PE) (clone C9B7W) | BioLegend | Cat#125208, RRID:AB_2133343 | B247016 |
| Rat anti-mouse CD267 (PE) (clone 8F10) | BioLegend | Cat#133404, RRID:AB_2240584 | B384620 |
| Rat anti-mouse CD274 (PE) (clone 10F.9G2) | BioLegend | Cat#124307, RRID:AB_2073557 | B223286; B246732 |
| Rat anti-mouse CD274 (APC) (clone 10F.9G2) | BioLegend | Cat#124312, RRID:AB_10612741 | B204731; B204732 |
| Rat anti-mouse CD279 (BV421) (clone 29F.1A12) | BioLegend | Cat#135217, RRID:AB_10900085 | B213656; B226274; B231473; B256184 |
| Rat anti-mouse CD366 (PE/Dazzle 594) (clone B8.2C12) | BioLegend | Cat#134013, RRID:AB_2632737 | B226637; B198857 |
| Rat anti-mouse F4/80 (PE-Cy7) (clone BM8) | BioLegend | Cat#123114, RRID: AB_893478 | B207313 |

| | | | |
|---|---|---|---|
| Rat anti-mouse FoxP3 (FITC) (clone FJK-16s) | eBiosciences | Cat#11-5773-82, RRID:AB_465243 | 2251241 |
| Rat anti-mouse I-A/I-E (BV510) (clone M5/114.15.2) | BioLegend | Cat#107635, RRID:AB_2561397 | B216155; B244226; B263357; B269686 |
| Rat anti-mouse IgM (BV510) (clone RMM-1) | BioLegend | Cat#406531, RRID:AB_2650758 | B230629; B272542 |
| Rat anti-mouse IgM (PE-Cy7) (clone RMM-1) | BioLegend | Cat#406513, RRID:AB_10642031 | B386037 |
| Rat anti-mouse Ly6C (PE-Cy7) (clone HK1.4) | BioLegend | Cat#128018, RRID: AB_1732082 | B357445 |
| Rat anti-mouse Ly6G (BV785) (clone 1A8) | BioLegend | Cat#127645, RRID: AB_2566317 | B366621 |
| Mouse anti-human Blimp-1 (AF647) (clone 6D3) | BD Biosciences | Cat#565002 | 1096352 |
| Mouse anti-human CD3 (APC-AF750) (clone UCHT1) | Beckman Coulter | Cat#A94680 | 200506; 200512 |
| Mouse anti-human CD4 (APC) (clone 13B8.2) | Beckman Coulter | Cat#IM2468U | 200092 |
| Mouse anti-human CD8 (KrO) (clone B9.11) | Beckman Coulter | Cat#B00067 | 200047; 200048 |
| Mouse anti-human CD19 (APC) (clone J3.119) | Beckman Coulter | Cat#IM2470, RRID:AB_130789 | 200050; 200071; 200072; 200076; 200085; 200096; 200097 |
| Mouse anti-human CD19 (PB) (clone J3-119) | Beckman Coulter | Cat#A86355 | 200025 |
| Mouse anti-human CD38 (FITC) (clone HIT2) | BD Biosciences | Cat#555459, RRID:AB_395852 | 6328698; 8179642; 9140760 |
| Mouse anti-human CD45 (PB) (clone J33) | Beckman Coulter | Cat#A74763 | 200044; 200045; 200046; 200048; 200049 |
| Mouse anti-human CD45 (KrO) (clone J33) | Beckman Coulter | Cat#A96416 RRID: AB_2888654 | 200110 |
| Mouse anti-human CD138 (PE) (clone PE) | Beckman Coulter | Cat#A54190 | 200031; 200037; 200045; 200046 |
| Mouse anti-human CD274 (PE-Cy7) (PDL1.3.1) | Beckman Coulter | Cat#A78884 | 200017; 200036; 200038 |
| Mouse anti-human CD279 (APC) (clone EH12.2H7) | BioLegend | Cat#329908, RRID:AB_940475 | B25407; B292213 |

| | | | |
|---|---|---|---|
| Mouse anti-human IgM (RB780) (clone G20-127) | BD Biosciences | Cat#569135 | 3027595 |
| Ultra-LEAF™ Purified anti-mouse CD274 (clone 10F.9G2) | BioLegend | Cat# 124318 | B192487 B390539 |
| Ultra-LEAF™ Purified Rat IgG2b, kappa Isotype Ctrl | BioLegend | Cat# 400644 | B416074 |
| Bacterial and virus strains | | | |
| Biological samples | | | |
| Patient blood samples | Hospices Civils de Lyon, France | N/A | |
| Healthy volunteer blood samples | Etablissement Français du sang, France | N/A | |
| Chemicals, peptides, and recombinant proteins | | | |
| Bortezomib | Sigma-Aldrich | Cat#179324-69-7 | |
| Dimethyl sulfoxyde | Sigma-Aldrich | Cat#D2650 | |
| Phytohemagglutinin (PHA) | Thermo Fisher Scientific | Cat#R30852801 | |
| $OVA_{323-339}$/CFA Emulsion | Hooke laboratories | Cat#EK-0132 | 0106 |
| $OVA_{323-339}$ in tissue culture media | Hooke laboratories | Cat#DS-0141 | 102-190224F |
| Critical commercial assays | | | |
| CellTrace™ Far Red | Thermo Fisher Scientific | Cat#C34564 | |
| Click-iT™ EdU Alexa Fluor™ 488 Flow Cytometry Assay Kit | Thermo Fisher Scientific | Cat#C10420 | |
| EasySep™ Mouse CD138 Positive Selection Kit | STEMCELL Technologies | Cat#18957 | |
| EasySep™ Mouse B Cell Isolation Kit | STEMCELL Technologies | Cat#19854A | |
| EasySep™ Mouse T Cell Isolation Kit | STEMCELL Technologies | Cat#19851A | |
| eBioscience™ FoxP3/transcription factor staining buffer set | Thermo Fisher Scientific | Cat#0055-23 | |
| ELISA MAX™ Deluxe Set Mouse IL-10 | BioLegend | Cat#431414 | |

| | | |
|---|---|---|
| Human IFN-gamma DuoSet ELISA | Bio-Techne | Cat#DY285B |
| MACSprep™ Multiple Myeloma CD138 MicroBeads, human | Miltenyi Biotec | Cat#130-111-744 |
| Mouse IFN-gamma DuoSet ELISA | Bio-Techne | Cat#DY485 |
| nCounter® Mouse PanCancer Immune Profiling Panel | NanoString Technologies | Cat#XT-CSO-MIP1-12 |
| PerFix-nc Kit (no centrifuge assay Kit) | Beckman Coulter | Cat#B31167 |
| Propidium iodide solution | Sigma-Aldrich | Cat#P4864 |
| RNA Lysis Buffer | ZYMO RESEARCH | Cat#R1060-1-100 |
| RNeasy Plus Mini Kit | QIAGEN | Cat#74134 |
| RosetteSep™ Human B Cell Enrichment Cocktail | STEMCELL Technologies | Cat#15024 |
| RosetteSep™ Human T Cell Enrichment Cocktail | STEMCELL Technologies | Cat# 15021 |
| Tag-it Violet™ Proliferation and Cell Tracking Dye | BioLegend | Cat#425101 |
| T Cell Activation/Expansion Kit, mouse | Miltenyi Biotec | Cat#130-093-627 |
| T Cell Activation/Expansion Kit, human | Miltenyi Biotec | Cat#130-091-441 |
| Zombie Aqua™ Fixable Viability Kit | BioLegend | Cat#423101 |
| Deposited data | | |
| Experimental models: Cell lines | | |
| Experimental models: Organisms/strains | | |
| Mouse: JAX™ C57BL/6J | Charles River | JAX: 000664 |
| Mouse: B6(Cg)-Il10tm1.1Karp/J | The Jackson Laboratory | RRID:IMSR_JAX:014530 |
| Oligonucleotides | | |
| Recombinant DNA | | |
| Software and algorithms | | |
| FlowJo software (version 10.8.1) | BD Biosciences | https://www.flowjo.com/ |
| Gen5 software 3.04 | BioTeck | https://www.biotek.com |

| | | |
|---|---|---|
| GraphPad Prism (version 9.0.2) | GraphPad Software | https://www.graphpad.com |
| Ingenuity Pathway Analysis | QIAGEN | https://digitalinsights.qiagen.com |
| Kaluza Analysis (version 2.1) | Beckman Coulter | https://www.beckman.com |
| R (version 3.6.2) | R foundation | https://cran.r-project.org/ |
| Other | | |
